# Experiences of health care from stroke survivors and caregivers from minoritised ethnicities

**DOI:** 10.64898/2025.12.15.25342150

**Authors:** Andrea Kusec, Xu Wang, Lindsey Thiel

**Affiliations:** Postdoctoral Research Associate, University of Oxford, Oxford, UK; Senior Lecturer in Psychology, Leeds Beckett University, Leeds, UK; Senior Lecturer in Speech and Language Therapy, Leeds Beckett University, Leeds, UK

**Keywords:** healthcare, stroke, ethnic minorities, health inequalities, culture, minority health

## Abstract

**Introduction:** Stroke disproportionately affects minoritised ethnicities. While quantitative evidence has shown a difference in stroke risk and type of care received between UK ethnicity groups, qualitative data is sparse. We sought to explore experiences of in-hospital and community-based care from stroke survivors and caregivers from minoritised ethnicities.DD

**Methods:** Audio recorded semi-structured interviews were conducted with stroke survivors and caregivers who self-identified as a minoritised ethnicity (e.g., Black, South Asian). Interviews covered experiences of incorporating cultural, religious, and/or dietary needs into stroke care, whether they perceived care was affected by ethnicity or cultural background, and ways to make care more culturally inclusive. Interviews were transcribed verbatim and analysed using reflexive thematic analysis.

**Results:** Twenty-four participants (n=16 stroke survivors, n=8 caregivers) took part. Themes included feeling different from a “typical” stroke survivor and affinity with British cultural norms (“*I Feel Different in Stroke Care*”); valuing culturally inclusive care but not always receiving it (“*Culturally Inclusive Stroke Care is Important but Inconsistent*”); individual perceptions of whether ethnicity affected care (“*Personal Interpretations of the Role of Ethnicity in Stroke Care”*); and tensions between caregivers advocating for cultural needs versus community perspectives of stroke (“*Families Champion Stroke Survivors’ Cultural Needs, What about the Community?”*).

**Conclusions:** Stroke survivors from UK minoritised ethnicity groups may feel “out of place” in care and may not receive sufficient cultural support. Individual interpretations of ethnicity, and affinity to British culture, affected perspectives on stroke care. Further efforts should be made to include culture and religion within person-centred stroke care.DD

## Introduction

Globally, stroke is the leading cause of lifelong disability [1], with the incident number of strokes expected to increase by twofold by 2050 [2]. This dramatic increase in stroke represents an aging population, but also improvements in medical care leading to fewer deaths post-stroke compared to previous birth cohorts.

Black and South Asian individuals are more likely to have a stroke compared to White individuals [3,4], and are the most likely groups to have a stroke in the UK [4, 5]. Differences in risk have ranged from 15% [3] to 400% [6]. Stroke is more likely to occur at a younger age in Black and South Asian adults [7, 8]. This increased stroke risk has been reported in North America [3, 6], the UK [9, 4], and India [10].

This differential risk has been linked to biological and psychosocial factors. Diabetes, elevated blood pressure, heart disease, and atrial fibrillation, all established stroke risk factors, have a higher incidence in Black and South Asian adults [9, 11, 6]. However, it is well documented that minoritised ethnicity groups in Western countries endure racial discrimination and disparities in quality and provision of stroke care [12, 13]. For example, Black stroke survivors in the UK and US have better post-stroke survival rates but worse functional outcomes that are not explained by demographic or stroke-related biological factors [14, 15].

While quantitative evidence is robust for stroke risk, quality of care received, treatments offered, and provision of care, there has been limited research on understanding lived experiences of healthcare from minoritised stroke survivors, particularly in the UK. Stroke survivors from minoritised ethnicities are underrepresented in research across a variety of settings (e.g., clinical trials) [16]. There have been several studies from the caregiver’s perspective; for example, the caregiver’s role in accessing long-term care [17], importance of trust and communication with social care workers [18], and supporting family involvement in stroke survivor’s care [19]. Qualitative research has examined professionals’ perspectives of working with stroke survivors from minoritised backgrounds – for example, professional capacity to integrate cultural considerations into stroke care [20], and challenges experienced by community nurses in providing home care to patients from marginalised ethnicities [21].

In a recent scoping review of post-stroke experiences and needs of South Asian communities in high income countries, Kokorelias et al. [22] identified only four qualitative studies exploring perspectives of stroke survivors. One study took place in New Zealand [23], two in Canada [24, 25], and one in the UK [26]. Key factors included the importance of within-community support, managing stigma surrounding their post-stroke symptoms, and needing to make substantial lifestyle changes, particularly around their diet and cultural expectations around returning to work. Similarly, Singh et al [27] report only two studies conducted in the UK with Black female stroke survivors, interviewing a total of 13 stroke survivors across two studies [28, 29]. Common themes included the importance of religion and spirituality toward stroke recovery, the importance of social networks, and adopting a positive mindset. More recently, Livingstone et al. [30] reported that Black stroke survivors in the UK report multiple barriers to navigating healthcare, including limited awareness of stroke risk and symptoms, not being listened to or believed by healthcare professionals, and misalignment of care with their cultural and age-related needs.

In summary, Black and South Asian individuals are more likely to have a stroke and at a younger age than White individuals. There is evidence of inequities in access to healthcare, interventions received, and outcomes in Western countries. There has been limited qualitative research exploring the perspectives of people across a range of minoritised ethnicities of their stroke care, especially from the stroke survivor perspective. There is a need to understand lived experiences and which aspects of care are considered to be important, so that these insights can support the planning of more equitable health and social care services.

### Study Aim

*What are the experiences of stroke survivors and caregivers from minoritised ethnicities of accessing and receiving UK health and social care services?*

## Methods

### Study Design

A qualitative design was employed to address the research aims. Constructivism was the underpinning theoretical framework as we sought to explore participants’ subjective realities which were socially and experientially constructed 31-33]. We aimed to understand insights from participants’ lived experiences of stroke-related health and social care through the lens of minoritised ethnicity communities.

### Study Sample

Participants comprised stroke survivors and informal caregivers of a stroke survivor (hereafter referred to as ‘caregivers’). An informal caregiver was defined as individuals who are unpaid and care for another person due to disability and/or illness [34].

Inclusion criteria for both stroke survivors and the caregivers were: (1) age ≥18 years; (2) being a UK resident living in the community; (3) able to speak and understand conversational English; and (4) self-identify as a minoritised ethnicity group (Gov UK, 2021). Additionally, stroke survivor participants were required to have had ≥1 stroke at least 3 months prior to the study. Caregivers were required to self-identify as someone who cares for a stroke survivor and could include spouses, parents, or friends, but excluded paid caregivers.

### Recruitment

Participants were recruited using purposive and convenience sampling. To maximise recruitment several methods were used, including (1) social media advertisement via X (formally Twitter); (2) UK stroke groups’ social media pages / WhatsApp groups with permission (e.g., UK Different Strokes Facebook page, Stroke User Group at Leeds Beckett University); (3) Newspapers, Newsletters and local radio (e.g., Yorkshire Evening Post, BBC Leeds, and Stroke News); (4) the Leeds Beckett University Speech and Language Therapy Clinic, and (5) our existing network of stroke survivors/caregivers who had participated in previous projects led by the authors. Through all methods, participation was emphasised as voluntary.

Following an expression of interest and eligibility screen, the researchers obtained informed consent over email or via an individualised QualtricsXM link. Each participant was provided a £20 high-street shopping voucher as a means of acknowledging their time and input.

### Data Collection and Study Materials

Semi-structured interviews were conducted via Microsoft Teams, telephone, or in person at Leeds Beckett University based on participant preference. Some stroke participants requested to participate in the interview with their caregivers to support communication difficulties.

Interviews (*Appendix 1*) consisted of open-ended questions with prompts to encourage insight of participants experiences of accessing and receiving health and social care when they were in hospitals and after being discharged to the community. Probes included how cultural, religious and dietary needs were incorporated into care, instances where they were made aware of their own ethnicity, and ways to improve cultural inclusivity in care. All interviews were audio-recorded with participant consent.

Participants provided demographic information including age, gender, ethnicity, time since stroke, years of education, occupation, and marital status. Caregivers were additionally reported number of years of caring for, and their relationship to, the stroke survivor. Stroke participants completed the 10-item Barthel Activities of Daily Living Index [35] to assess functional ability. The Barthel ranges from 0 (more severe disability) to 20 (greater functional ability).

### Positionality

SXW is an able-bodied senior lecturer in health and clinical psychology. XW has worked in the context of psychological support for stroke survivors and family caregivers, and secondary prevention for younger strokes. Coming from a minoritised ethnicity group herself, XW is deeply motivated to hear participant’s voices in this research. XW views community support as equally important to family support for stroke survivors, and her view of the data was influenced by both this and her research expertise with younger stroke survivors.

AK is a mixed (White South Slavic and Mestizo) ethnicity, able-bodied postdoctoral researcher in neuropsychology. AK has worked with a range of stroke and acquired brain injury survivors with a range of backgrounds and post-brain injury profiles in clinical research settings. AK’s research experience with stroke survivors has been in the context of mental health and cognition-based intervention research both acutely and in the community, giving her an understanding of the care pathway post-stroke. AK views the experience of post-stroke disability to have an intersectional impact with pre-stroke systemic inequalities including due to race and ethnicity when engaging with health care services, and her view of the data was influenced by both this and her psychology research background.

LT is a white, British, able-bodied Senior Lecturer in Speech and Language Therapy. She has worked with stroke survivors with language and communication difficulties in clinical practice and research; she was therefore well-suited to interview stroke survivors with aphasia. She used supportive conversation techniques so that participants could share their experiences within interviews. Participants were encouraged to provide answers through talking, but also writing (in the Teams chat), and gesturing. LT teaches the importance of promoting bilingualism and equitable healthcare within her role and acknowledges that her interpretation of the data has been influenced by this.

### Data Analysis

Interviews were transcribed verbatim by two postgraduate research assistants. Comments on non-verbal methods of communication (for example gesture and writing) were also transcribed.

Reflexive thematic analysis (RTA) was used by AK, SXW and LT to analyse the data, involving the generation of codes and clearly defined themes to make sense of participants’ experiences [36] (Table 1). Analyses were conducted using Microsoft Word (coding transcripts) and Microsoft Excel (code refinement, candidate theme generation, and theme refinement and finalisation).

**Table 1.** Research Team Reflexive Thematic Analysis Procedure.

| **Reflexive Thematic Analysis Phase** | **Research Team Corresponding action** |
| --- | --- |
| Phase 1: Familiarisation with the data | Transcripts were read and re-read by AK, SW and LT and initial ideas were noted. |
| Phase 2: Generating initial codes | The data were coded inductively, by working through each transcript and identifying both semantic and latent meanings. Coding was carried out using a manual process, where codes and reflexive comments were typed into a column alongside the data within a word processing document. AK coded the entire data set while LT and SW each coded half of the data set. Codes were shared and discussed during online meetings. |
| Phase 3: Constructing themes | Each research team member individually sorted codes by examining them for patterns, with any that were related in meaning being organised into lists of candidate themes and subthemes. The researchers met to compare candidate themes, and through in-depth discussion reduced themes to those that were common across the three analysts and captured distinctiveness of experiences to best address the research question. |
| Phase 4: Reviewing themes | Several meetings were conducted to iteratively refine the themes and subthemes while reviewing and considering whether they reflected the coded data extracts and the data set as a whole. Some candidate themes were merged or divided. |
| Phase 5: Defining and naming the themes | The themes were divided among the researchers, and each wrote a summary to understand the essence of each theme. These were shared and presented within meetings to generate further discussion and to make final changes to themes. |
| Phase 6: Producing the report | This phase included the final analysis and write-up by all members of the research team. Anonymised extracts from the data were selected to demonstrate the themes and subthemes. The research team shared themes with a charity group leader who is a caregiver of an ethnic minority stroke survivor. They |
|  | discussed the themes and were given feedback to ensure their clarity and comprehensiveness. They suggested expanding on elements of British-born vs immigrant participants from minoritised backgrounds, how different ethnicity groups may interpret their experiences, the pressure of assimilating to a colonial White norm of productivity, and within-community stigma of disability, being viewed as a disabled person, and how this interacts with engaging with their community post-stroke. This feedback was incorporated when writing the report. |

### Rigour

The researchers engaged in continuous reflexivity through keeping their own reflexive notes, and through discussions within meetings. Authors were aware that they all had unique perspectives of the data, and this was seen as a strength in developing themes. The researchers immersed themselves in the data, leading to an in-depth and comprehensive analytical process. A detailed audit trail was kept throughout the stages of analysis.

## Results

Twenty-four participants (16 stroke survivors, 8 caregivers) took part (Tables 2 and 3). Interviews took approximately 45 minutes each (minimum 19, maximum 74 minutes).

**Table 2.**
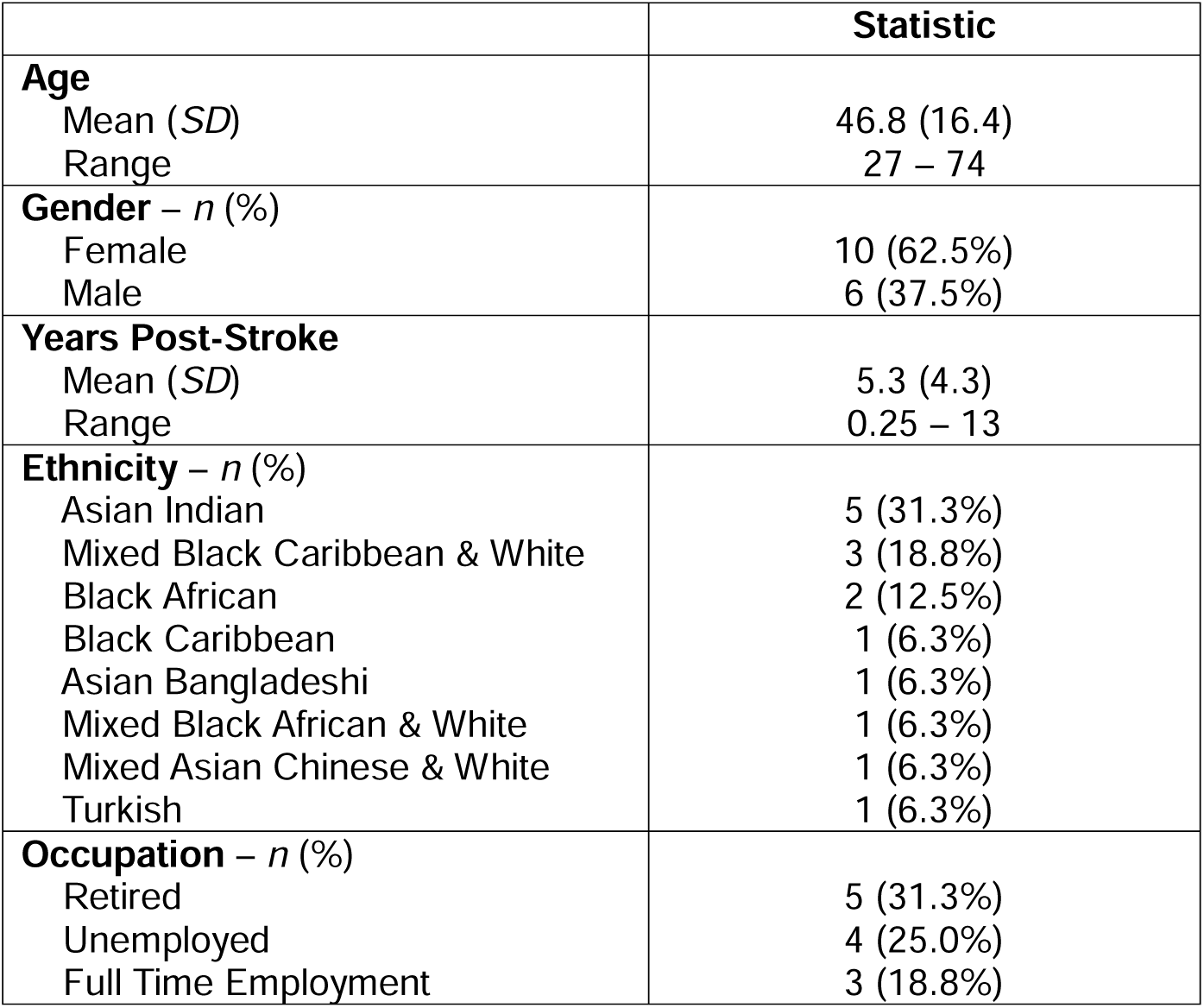
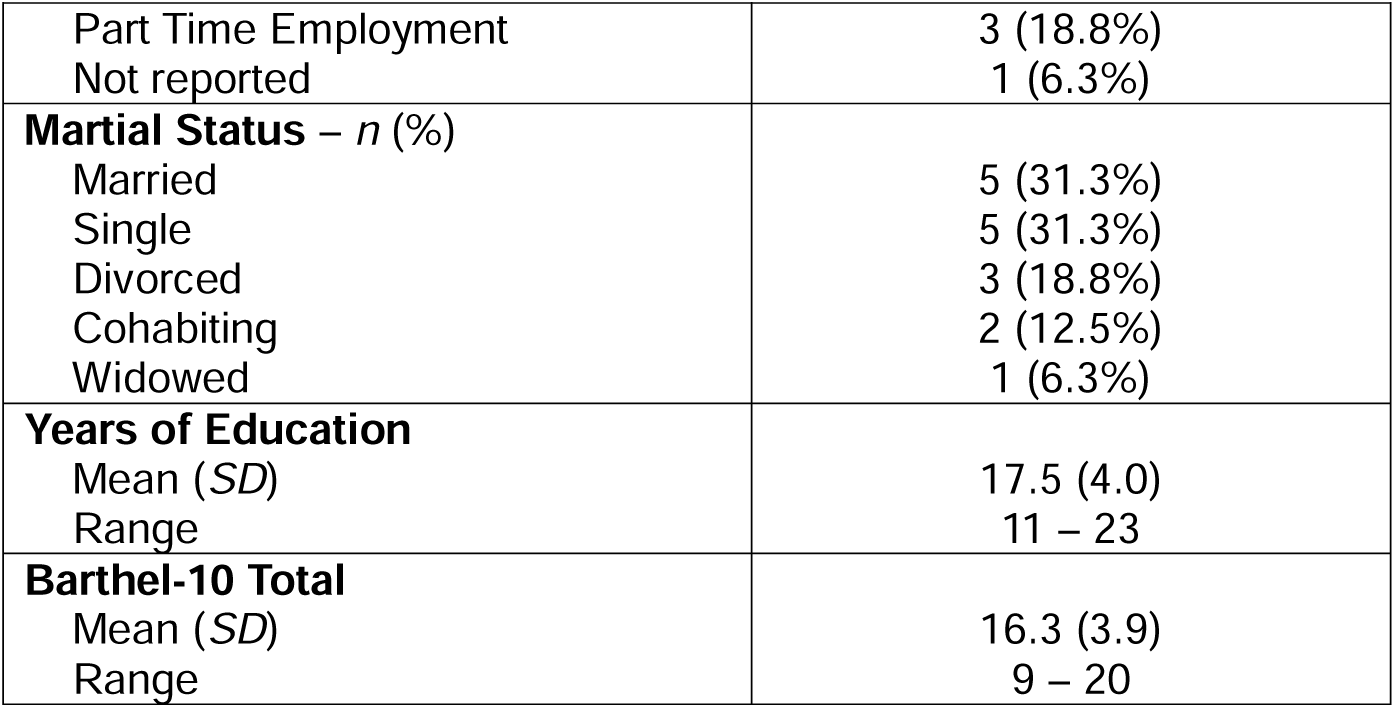
Demographic data for stroke survivor participants (n = 16).

**Table 3.** Demographic data for caregiver participants (n = 8).

|  | **Statistic** |
| --- | --- |
| **Age** |  |
| Mean (SD) | 46.3 (12.2) |
| Range | 28 – 63 |
| **Gender – n (%)** |  |
| Female | 4 (50.0%) |
| Male | 4 (50.0%) |
| **Years as Carer** |  |
| Mean (SD) | 3.3 (2.6) |
| Range | 1 – 9 |
| **Ethnicity – n (%)** |  |
| Asian Pakistani | 3 (37.5%) |
| Black Caribbean | 2 (25.0%) |
| Asian Indian | 1 (12.5%) |
| Asian Chinese | 1 (12.5%) |
| Mixed Black Caribbean & White | 1 (12.5%) |
| **Marital Status – n (%)** |  |
| Single | 4 (50.0%) |
| Married | 3 (37.5%) |
| Cohabiting | 1 (12.5%) |
| **Relationship to Stroke Survivor – n (%)** |  |
| Son | 4 (50.0%) |
| Daughter | 2 (25.0%) |
| Married Partner | 2 (25.0%) |
| **Years of Education** |  |
| Mean (SD) | 17.4 (3.4) |
| Range | 11 – 22 |

Four broad themes were constructed (Figure 1). *Supplementary Table 1* shows example quotations for each theme and subthemes.

**Figure 1.**
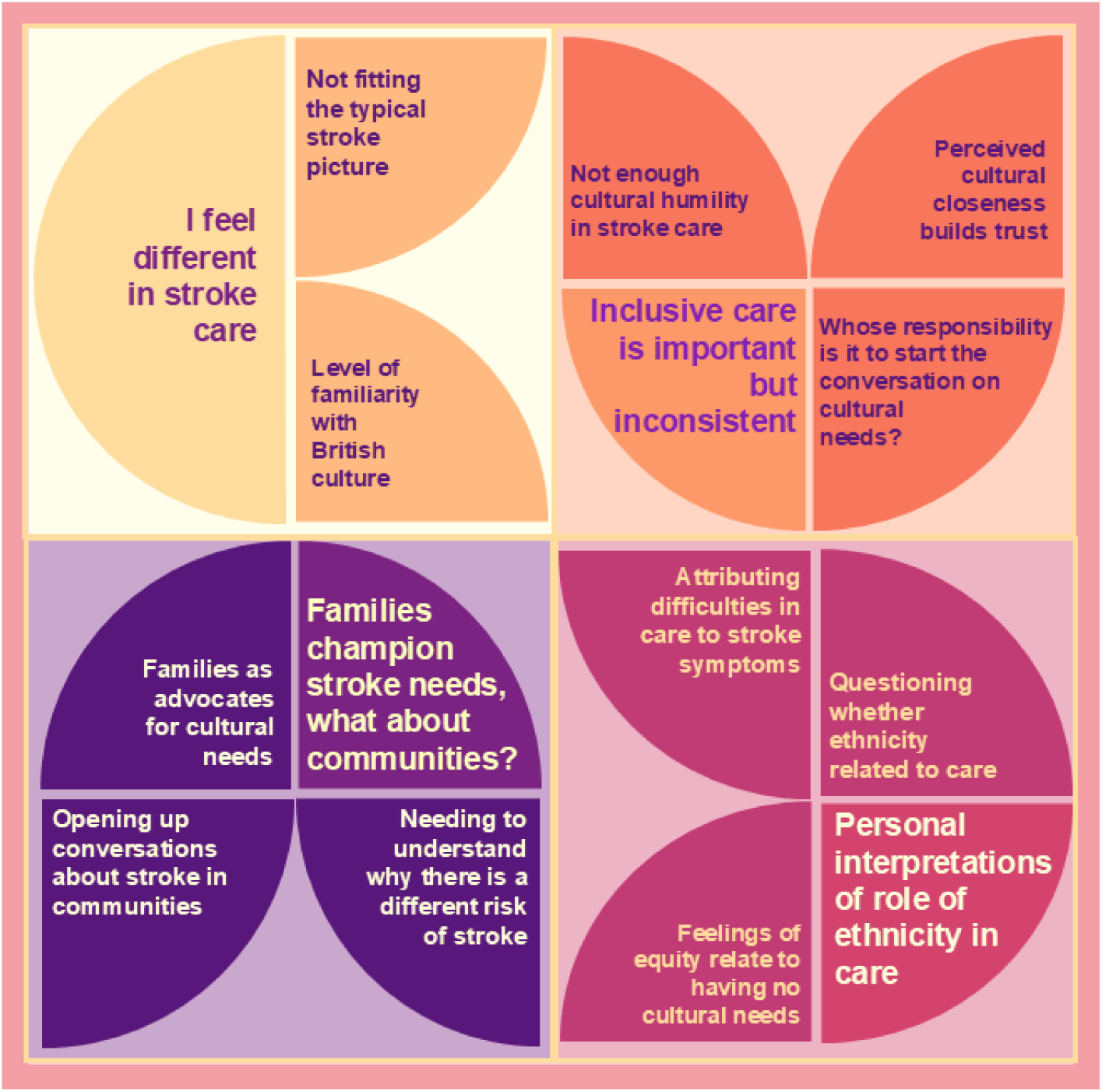
Visualisation of the final constructed framework. Each quadrant represents an overarching theme with corresponding subthemes, with the overarching theme in larger text.

### Theme 1: I feel different in stroke care

Participants felt different from others throughout their stroke care process, from diagnosis, hospitalisation, and community care. This impression was tied to feeling their stroke diverged from a typical stroke profile and not often meeting other stroke survivors from their own ethnicity group and their affinity with British culture.

#### Subtheme 1: Not Fitting the Typical Stroke Picture

Participants often expressed that they thought stroke mainly happened to “*old White people*” which was viewed to be the “typical” stroke profile. Participants felt different from other stroke survivors in their ethnicity, age, native language, or a combination of these elements. For example, a young stroke survivor said:

> *“When [healthcare professionals] told me I was having a stroke, [I] didn’t believe it because that’s something old white people have, and that’s not me.”* (P8, stroke survivor)

Similar experiences of feeling “othered” were reported by other stroke survivors. For example, when using an online stroke peer support forum, one participant noted:

> *“When I talk to people on the [online stroke peer support board], they don’t know what colour I am…then they meet me and they’re like ‘Oh, hang on, like…’ and I can see it. They’re not saying it, obviously, but they can go ‘Oh, but you’re not, you’re not white.’ [chuckles] Like, ‘Well, why would I be?’ Like, you know, we live in a multicultural society.”* (P23, stroke survivor)

Other ways that participants expressed feeling different was by having rare forms of stroke that do not align with typical “stroke risk factors” like poor diet and lifestyle factors. From this feeling of “not fitting in”, participants reported the desire and need to find stroke survivors similar to them throughout their recovery process, both in terms of age, ethnicity, language, and/or stroke profile. Participants described engaging with group-based therapy or peer support groups that were attended by predominantly White or British stroke survivors in an attempt to achieve this.

> *“We all go through this [stroke recovery] journey alone…In the journey through my health care in all of these hospitals, the health professionals see hundreds of us, and the one thing that I have said consistently that was missing and lacking is the ability to see, meet, share, empathise, talk through stories, of our journeys and healing process. And it feels incredibly frustrating to me that, that wasn’t, and isn’t there. And people in the Black and South Asian community are often in large families. And, and a community approach, or a group approach, I think would be so beneficial…I can tell you today, what, 18, 20 months after my stroke, I haven’t met anybody who is like me. Who is also non-White, who has had a stroke. And I think that speaks volumes.”* (P24, stroke survivor)

#### Subtheme 2: Level of Familiarity with British Culture

Participants referenced how their level of familiarity with British culture and the UK health care system affected their stroke care experiences. A greater closeness to British culture was sometimes thought as beneficial and reduced feelings of being “different”.

> *“I think if I was, if I hadn’t married someone that’s British, I would be very, uh, foreign to the culture here. So, in hospital, I definitely would feel, uh, more isolated, and more different than everyone else”* (P8, stroke survivor)

Stroke participants who were born or have lived for a long time in the UK similarly described themselves as being predominantly “British” and not feeling different:

> *“…mainly because I’ve been here for 50 years, I, I consider myself more British than Asian.”* (P18, stroke survivor)

Conversely, some participants who felt less familiar with British cultural norms found this to cause “feeling different.” For example, a participant who immigrated to the UK in adulthood, described their hospital experience as “*alienating*” and care received as “*straightforward*”:

> *“If you’re in an atmosphere that’s everybody’s born and raised here, and you’re just like came afterwards, there’s a bit of feel, alienation feeling.”*

> *“…because people were so straightforward, uh, I, I was even having, thinking twice before I was call, calling the nurse to ask for something…If I was in [home country], they would be basically, bit more, uh, you know, warm-hearted. But I’m not saying it in a rude way…it’s their own culture…but when you come from a culture like that, especially when you’re sick, it just kind of makes you feel a bit more shy.”* (P7, stroke survivor)

### Theme 2: Culturally inclusive stroke care is important but inconsistent

Participants reflected on how their culture, ethnicity and religion were acknowledged, considered and incorporated. While some noted that there were healthcare professionals who acknowledged their religion and faith in care, others felt that it was crucial for professionals to provide individualised care that met their cultural and religious needs. Despite participants emphasising the necessity for culturally inclusive support, this was not consistently provided.

#### Subtheme 1: Not enough cultural humility in standard stroke care

Participants described a lack of acknowledgement from professionals about their culture, and in some cases, assumptions were made about their needs. One participant described how she had experienced discrimination towards her aunt’s religious beliefs of physical and spiritual cleanliness. For Muslims to have physical cleanliness requires washing themselves with water:

> *“We knew that my aunt would want to be just washed with a trickle of water and that was it, whereas the nurses said, ‘oh, we can’t do that, we can’t do that.’…‘if [the nurses] were to take her out to the toilet, we’re just going to let her sit on the toilet and a dry tissue will be used’… for us that’s not in along the rules of [Islamic] cleanliness…it’s not acceptable according to how we cleanse, just where the trickle of water running off the body, and that’s fine.”* (P14, caregiver)

Another aspect discussed was the traditionally English food served in hospitals, with some participants not being offered alternatives. Sometimes, assumptions were made by staff about what a stroke survivor might want to eat:

> *“When my son went to the hospital, he, he said ‘He never got fed.’ [The nurse] said, ‘Well, we gave him the Indian food and everything, right?’. And then [my son] says, ‘He won’t eat that Indian food’, because I know I won’t eat it. He knows I won’t eat it.”* (P19, stroke survivor).

Multilingual stroke survivors often only received speech and language therapy in English, which was particularly problematic within speech and language therapy services as described by one participant:

> *“I’m not saying they everyone needs to be from different backgrounds and, and you need to have speech therapists that relates solely to your heritage. But, perhaps a mix or a better mix would be useful…for my dad it, it, it would have been useful to help him develop his Gujarati. It’s not bad, but it’s no way near the same level as his English.”* (P11, Caregiver)

In contrast, some participants described positive experiences of culturally or religiously inclusive support received, such as opportunities to meet people of the same faith in their stroke support group. Similarly, other participants reported positive interactions with staff around understanding the importance of religious beliefs, being offered Halal food, and celebrating holidays like Eid:

> *“[My doctor] understood my, um, religion and my faith and I think we have the same feeling, and that way he was able to understand, erm, the strength of belief because that was the only realistic thing I was holding onto for a long time.”* (P2, stroke survivor)

#### Subtheme 2: Perceived cultural closeness builds trust

Participants discussed feelings of closeness when they felt their cultural needs were acknowledged and addressed by healthcare professionals, enhancing their sense of trust. Sometimes, the source of comfort and closeness was from healthcare professionals who were also from marginalised ethnicities:

> *“I remember the specialist being quite kind and quite careful. She was an ethic minority herself as well, so maybe that made me feel slightly more comfortable”* (P9, stroke survivor).

Participants suggested that, in their experience, healthcare professionals from minoritised ethnicities were viewed to have more cultural humility, empathy, familiarity with different cultures, and understanding due to their own diverse backgrounds. This was not exclusive to ethnicity, and could extend to being part of a non-majority background:

> *“The [domiciliary carers] were foreign, they, even the Portuguese lady, who, while being White, still seemed to have more of an awareness in a sense, as to what was needed in a sense or, or, or how we would do things”* (P22, caregiver)

Other participants described positive experiences relating to being cared for by someone with a similar race, ethnicity or culture.

> *“The first assistant that came was an African lady […] I think it was Uganda. Maybe. So, she had some sort of a sense of, you know, what Black people, what Afro-Caribbean people would need…I guess you could call it a cultural understanding […] it was implicit almost.”* (P022, caregiver)

The consequence of being cared for by somebody that participants did not feel any cultural closeness to was a lack of connection, and even a lack of trust, in the professional’s ability to carry out their role effectively.

> *“You really don’t want to get a White caregiver or somebody who that they won’t feel connected to take care of their personal needs. Because the thing that would happen is, the healthcare plan wouldn’t get used because, um, there’s that missed connection that’s not there, and the caregiver wouldn’t be able to effectively achieve their role because the patient doesn’t feel that trust, that connection, and would not be able to request for some care from the caregiver.”* (P01, Caregiver)

#### Subtheme 3: Whose responsibility is it to start the conversation about cultural and religious needs?

Participants felt uncertain about who should begin the conversation about cultural and religious needs in healthcare settings. Having cultural or religious needs different from the norm was dependent on the stroke survivor or a family member starting this conversation. Some felt that healthcare professionals should be asking about any cultural, religious, or dietary needs, because if they did not share anything about their own needs then they would not be asked.

> *“Unless you say ‘well I’m say a Muslim, or, or Hindu, and I’m a vegetarian’ and so forth, unless you kind of outwardly present, then those things are not going to be asked.”* (P22, caregiver)

Healthcare professionals exploring their clients’ cultural needs was viewed to be part of having an inclusion and diversity mindset. A key aspect of this was active listening to client preferences. Some participants felt that professionals should be regularly learning about different cultures and cultural customs and be responsible for beginning the conversation on cultural needs. For example:

> *“I think the best thing would be for the staff to have refreshers [on patients’ cultural/religious needs] as often as possible…they need to have a bit of background, cultural background, about those religions and faiths and backgrounds and, and cultures and all that before they start handling those patients or going to their houses…So like for example, you know, umm, nursing staff, people who came, you know, cause it’s not always easy for, for people who come into your house to ask them, ‘Oh, could you please take your shoes off before you come inside?’”* (P15, Caregiver).

### Theme 3 – Personal interpretations of the role of ethnicity in care

When reflecting on both positive and negative experiences of stroke care, participants attributed these to different causes. Some began questioning and wondering whether they were treated differently or received inadequate care because of their ethnicity. Others believed that any difficulties accessing care were due to their stroke symptoms, such as aphasia, rather than their ethnicity. Participants who felt they had no cultural or religious needs different from what is routinely provided stated feeling treated equally.

#### Subtheme 1 – Questioning whether ethnicity related to care

When looking back at their experiences, some participants noted that as time passed, they started considering whether their ethnicity did affect care decisions made. Participants were often unable to make definitive statements about this, and instead noted its ambiguous role:

> *“I do wonder more and more some of the decisions and ways in which I’m prioritised or not prioritised may in at least partly be due to my ethnic background.”* (P9, stroke survivor)

This ongoing questioning led to reflections across a range of elements of health care, for example from the number of physiotherapy sessions offered, decisions on prioritising return to driving, and general follow up and care offered:

> *“At times, to be honest with you, I end up at times thinking, is it, is it a mistake that I’m Black? ‘Cause most of the times, for example like right now, I was given, they say they can’t do any more, like, on the physiotherapy. Yes, I can walk a bit…but I don’t know, I don’t know.”* (P17, stroke survivor)

Others reflected on the possibility that ethnicity could regularly be affecting care, but a lack of awareness of what cultural support would be available to them could lead stroke survivors to perceive there to be sufficient support:

> *“Sometimes we find that, uh, a service, may lack some element…sometimes the recipient isn’t aware of what could have been available or what could have been asked, um, or what could have been offered, and didn’t even realise, let’s just say they didn’t even realise they were being discriminated against, or they didn’t realise that there was a, there was a prayer room, you know, whatever it may be…So they never thought of themselves as having received substandard care”* (P024, Stroke Survivor).

#### Subtheme 2 – Attributing difficulties in care to stroke symptoms

For some, any perceived gaps in care were attributed to their newly-acquired stroke symptoms, chiefly speech and language or cognitive changes:

> *“I don’t look at it on the minority or anything, I just look at the disability. In the stroke, it’s people, like people who are not feeling well, mentally or anything.”* (P017, Stroke Survivor)

Some participants disregarded the role of ethnicity completely, stating that the emotional and physical impact of the stroke itself was more important:

> *“I don’t think [I was made aware of my ethnicity]…I was more heartbroken and hurt regarding the brain bleed and spending my time over in [the hospital].”* (P006, Stroke Survivor)

> *“I think the stroke issues are more human body related….and that, that had nothing to do with, uh, any mentions about religion or ethnicity. I just feel that the fact that I’m, I’m Indian, doesn’t make my body any different to an English man’s body.”* (P18, Stroke survivor)

#### Subtheme 3 – Feelings of equity relate to having no cultural needs

Some participants viewed themselves as having no cultural, religious or ethnicity-based needs that were different from the perceived “norm” of UK stroke healthcare:

> *“Because I suppose, I’m mixed race, and, umm, I don’t really have any kind of religion. I’m just, you know, treated normally…So, I can’t really, umm, answer that one because I have no, my experience of just, quite straightforward.”* (P006, Stroke survivor)

These participants perceived their needs, or their level of proficiency with the English language, to be similar to the majority of the population within the UK:

> *“Dad, do you think that your, your experience and how you related to the speech therapist would have been affected if they were Gujarati or not?”* (P011, Caregiver of P010)

> *“No, no. That don’t matter because I speak English language very well.”* (P010, stroke survivor)

Finally, some perceived there to be no differences in their care and their overall experience was a feeling of equality:

> *“I just don’t think they look at me in that way. They just look at me as a person, that’s had a stroke.”* (P006)

### Theme 4: Families champion stroke survivors’ cultural needs, what about communities?

Both stroke survivors and caregivers discussed interactions with family and the community in their stroke recovery journey. This theme recognises the tension between caregivers and family members often advocating for stroke survivor needs, but stigma around stroke and disability in the community persist.

#### Subtheme 1: Families are advocates for stroke survivors’ cultural needs

Many participants noted how families were devoted to caring for stroke survivors both in hospital and at home, often pushing for appropriate cultural, religious, and dietary needs of the stroke survivor to be addressed, such as chaplaincy services:

> *“I knew for my aunt that [chaplaincy services] was an absolute want, a desire, a need, a necessity…So maybe day one, it wasn’t a priority, but it was on day three. And then day four, my priority was on washing, bathing, cleaning and then maybe day seven, we were trying to recall, revisit chaplaincy services again, you know, so I was on top of that for all of this diary management of needs. But why were the healthcare assistants not? Why were nurses not?”* (P14, Caregiver)

Family members were often involved in supporting dietary preferences. This included respecting food preferences as part of their heritage, but also not assuming that if a stroke survivor was Indian that their preference was for Indian food:

> *“[My dad] got the same [traditional British food] every single day. It was only when I, or, or my friends, uh or relatives brought food into the hospital was he given the food actually wanted.“* (P11, caregiver)

#### Subtheme 2: Opening up conversations about stroke in minoritised ethnicity communities

Whilst cultural needs were actively advocated by families, participants often noted that stroke and disability are not openly discussed in their communities, potentially reflecting disability-related stigma:

> *“I can’t speak necessarily for other ethnicities and other minorities…I can say certainly among sort of Afro Caribbean’s or Caribbean people, they’re, discussing health issues is, is kind of taboo. It’s kind of ‘none of your business’”* (P22, Caregiver)

Some participants felt that this reluctance, or even resistance, to have an open conversation on health and stroke could be linked to a lack of awareness and knowledge of stroke, which in turn may lead to avoiding help from healthcare systems. When asked why people do not talk about stroke or disability openly in the community, participants discussed potential reasons:

> *“It’s a very close-knit community, they’ll celebrate, like weddings, and they’ll celebrate, you know, the son’s gone to university, or they’ll celebrate grandchildren. But anything that seems slightly negative, which a lot of medical issues unfortunately do, are, um, and especially when they can’t understand it…but the stroke, I think it’s because no one understands it as much…I don’t think a lot of these people probably get seen to, until it’s too late.”* (P023, stroke survivor).

When asked what could be done to promote open dialogue on health and stroke within communities, participants suggested conducting outreach work with community leaders, having younger generations who speak the same language communicate the impact of stroke, and using social and religious networks:

> *“We need the younger generation who understands [stroke or disability], who are like physios, the doctors and nurses, who can be involved to talk in their own language and explain to them…This is what you should be doing…if you speak in your own language, I think it breaks the barrier straight away.”* (P019, Stroke survivor).

#### Subtheme 3: Needing to understand why there is a different risk of stroke

Participants, particularly younger stroke survivors, reported a need to understand why people from minoritised ethnicities were at higher risk of stroke. For some, knowledge that minoritised ethnicity groups were at greater risk of stroke was not known prior to their stroke:

> *“One thing that interested me what you said, is that, um, you find that there’s more strokes that go on in the ethnic minority. And I wasn’t aware of that. I mean, to be fair, I never really experienced anybody that had struggled except for myself. So, um, I, I didn’t even know what was wrong with me.”* (P006, Stroke survivor)

Participants noted some potential reasons, but emphasised that understanding this disproportionate representation was important:

> *“The other question, the other challenge to some extent is, trying to find out why it is that we are disproportionately represented. I mean, do we, do we typically have higher cholesterol levels than, than the, the, the median group, I don’t know. Are we less active than other people? I don’t know, because typically you know, most Black people tend to have fairly active work lives at least.”* (P22, Caregiver)

Being provided with information that they are at a disproportionate risk of stroke, and being able to be told why they had a stroke, was viewed to be critical to post-stroke care:

> *“I question why this has happened to me, because everyone’s like ‘well, it’s your race.’ And I’m like, right, so I know cholesterol should be higher, but, I don’t have the same diet as my parents…So, I’m still struggling to understand why, where does that factor in our make-up of our body? And when it’s so about, ‘well, you’re*

> *Black’ or it’s like, you know…why is it such a big disparity between Asian, Black, ethnic, um, to the White person?…I think I get it if you’re in different countries because that lifestyle, that food, that weather, you know, that, there, there’s so many different contributing factors to that…But when you’re in the same country and you’re born here. And if it wasn’t for the colour of my skin and my name, and you’d think, you know, I was a White person. So um, what is it in my make-up, my genes, that’s caused this?”* (P023, Stroke Survivor)

## Discussion

This study found that stroke survivors from minoritised ethnicities are more likely to feel “othered” during stroke care and that culturally inclusive care is not consistently provided. Participants had differing interpretations of the role their ethnicity plays in care received. Family members were advocates for increased cultural support, but discussions about stroke within communities was viewed to be important.

Participants described feeling different from the typical stroke picture as someone with a minoritised background, especially younger stroke survivors. Although South Asian and Black individuals are the most likely groups to have a stroke [4, 5], they are still the minority [37]. Furthermore, minoritised ethnicity groups are underrepresented in the Sentinel Stroke National Audit Programme [38], suggesting a discrepancy between prevalence and service visibility. This finding of feeling othered reflects experiences reported in Livingstone et al. [30], where participants described feeling that, due to their ethnicity and age, they did not belong on stroke wards, and people did not look like them at peer support groups. These combined findings highlight the need to reduce “otherness” through increasing public awareness of stroke, reducing stigma, and providing healthcare professionals with information on stroke prevalence in these populations. Culturally inclusive community-led initiatives for stroke survivors from minoritised ethnicities (e.g., Different Strokes Black and Asian Stroke Survivors Project [39] should be established to support belonging and challenge systemic exclusion.

Participants stressed that culturally inclusive stroke care is essential, but lacking. This echoes previous research demonstrating racialised disparities in quality and provision of stroke care for minoritised ethnicity groups in Western countries [21, 30, 12, 13]. Contributing factors include professionals’ fear of making mistakes or causing offence, burnout, implicit bias, time constraints, inadequate support, lack of cultural humility training, scarcity of culturally and linguistically relevant materials, and insufficient collaboration with interpreters [21, 40, 41, 42, 43]. This results in efficiency being prioritised over equity, perpetuating White, mono-cultural, middle-class, Eurocentric norms.

Most participants were not asked about their cultural or religious needs by professionals, causing structural inequities for individuals with communication difficulties, limited English proficiency, or without a caregiver advocate. Participants’ desire for professionals to initiate these conversations supports research on the importance of open, sensitive communication, respect for cultural differences [44], empathy, curiosity, and responsibility to accommodate clients’ preferences [43]. Livingstone et al.’s [30] participants described distrust towards healthcare professionals due to past experiences of racism, discrimination and stereotyping, and that continuity of care, collaboration, and being given time to talk helped to build up trust.

When participants had their cultural and religious needs met, they trusted and feel closer to healthcare professionals, which happened more often with professionals from marginalised ethnicities or those with a shared cultural identity. This highlights the importance of racial and ethnic concordance [45], reflection on cultural values to increase cultural humility [44], the need for a diverse healthcare workforce [46], and having cultural humility be seen as essential to person-centred care [47].

Participants’ experiences of care were affected by their own interpretations of ethnicity in healthcare settings. For some, receiving inadequate care was attributed to their ethnicity. Singh et al. [27] note that Black stroke survivors in high income countries report distrust in healthcare providers more generally, which could extend to scepticism in whether they were provided appropriate cultural or religious care. Other studies note that healthcare providers may not fully understand minoritised ethnicity cultures [48, 19], negatively affecting care. Minoritised stroke survivors have reported feeling that rehabilitation teams work harder with White stroke survivors [49], aligning with participants’ views that disparities in their own care reflect general racial disparities in the UK.

Understanding whether ethnicity resulted in discriminatory practices was often met with uncertainty about whether this was due to poor stroke care more generally, or difficulties communicating needs given stroke-specific symptoms such as aphasia or disorientation. For example, healthcare professionals misunderstanding their speech, or the stroke survivor being unable to remember information. Many studies, both in minoritised ethnicities and in White stroke survivors, report feelings of stigmatisation and social isolation as a consequence of stroke [22]. Coping with new stroke symptoms may override the importance of ethnicity in care for some.

If there were no perceived cultural or religious needs, participants viewed themselves as being treated “normally.” A UK study of Black stroke survivors has found that some do not wish to engage with culture-specific or ethnicity-specific stroke healthcare programmes, as they have their cultural needs met elsewhere [19]. Possibly, the current UK standard of care already meets cultural and religious needs for some, but as indicated here, this may not be flexible enough to adapt to a wide range of needs.

Family members acted as champions for stroke survivor’s cultural needs across health care settings; however, this was not mirrored in responses that communities had toward stroke. This extends findings that caregivers often act as advocates more generally for stroke survivors [50]. It is possible that caregivers from minoritised ethnicities face additional logistic hurdles – in a health care system where cultural and religious needs are minimised or disregarded, caregivers will already face the increased responsibility of ensuring adequate care for their loved ones, but also have requests for additional religious, cultural or dietary needs dismissed [17]. Future work should consider both direct benefits to stroke survivors in improving inclusive care, but also indirect caregiver benefits.

Working with communities was viewed to be important to reduce stroke stigma, opening up discussion about stroke, understanding what a stroke was, and why minoritised ethnicity groups were overrepresented. Similarly, Livingstone et al. [30] noted that Black stroke survivors described not being aware that stroke is more prevalent and has a younger onset in Black communities, which in some cases led to participants not attending screening appointments. Within-community advocates were viewed to be valuable to have a shared common language and understanding of community-specific needs. In a review of interventions to increase stroke awareness, evidence in terms of effectiveness is lacking [51], with no common theoretical framework for intervention in minoritised ethnicity groups [52]. A combination of proximate determinants (e.g., health literacy interventions) and structural changes reducing healthcare inequalities can impact on awareness and stroke preparedness in at-risk communities [52].

### Clinical Implications

This study shows how structural inequities and systemic racism impact stroke care experiences. Our results highlight the importance of creating opportunities for individuals to connect with others who share similar cultural, linguistic, and religious identities. More awareness-raising projects about stroke within minoritised ethnicity communities. Healthcare professionals should start the conversation about cultural and religious needs, and include culture and religion into person-centred [53]. Reflecting on one’s own culture, values, power and privilege is an important starting point. Providing opportunities for patients to engage in cultural and religious customs and celebrations within their stroke care and to use their preferred language would improve inclusion and equity [53], and challenge the dominance of monolingual English norms [54]. Therapies should be delivered in the client’s preferred languages and interpreters used when necessary to prevent erosion of identity, relationships, and ability to engage in cultural and religious practices [55]. Family members should be actively involved in healthcare discussions and recognised as advocates [56]. For stroke care to become equitable, systemic change grounded in anti-racist policies and practices is needed [41].

### Strengths, Limitations and Future Directions

Our study included perspectives of stroke survivors and caregivers, creating a more holistic understanding of healthcare experiences in minoritised ethnicity groups. Our joint analysis process, including double coding across transcripts and several collective iterations of theme refinement, presents a thorough and transparent account of theme construction. Careful reflexivity was conducted, with an evidenced audit trail of decision making. Analyses were robust due to discussion with charity representatives working with a range of minoritised stroke survivors who additionally have lived experience of being a caregiver of a stroke survivor.

Limitations included a limited budget for translation services, though we have ample representation of stroke survivors with communication difficulties. Though our research team represents a range of backgrounds including minoritised and mixed ethnicities, the team did not have Black or South Asian researchers who could provide this additional lens to the data. Participants also had high levels of education and functional ability, which may limit conclusions made from stroke survivors with lower education levels and more severe strokes.

### Conclusion

Stroke survivors from minoritised ethnicity groups are at risk of “feeling different” and being othered when accessing healthcare. This may be amplified by the inconsistent nature of culturally inclusive stroke care, or whether they perceive their ethnicity to have played a role in care decisions made. Family members were often advocates for increased cultural support, but stigma around stroke was viewed to be important to discuss within communities. Future research should create partnerships between health care systems and communities to facilitate improvements on culturally inclusive care.

## Data Availability

Data are not available due to interview data containing potentially identifiable information.

## Acknowledgements

We would like to thank Janeil Bennett and Fatimah Bint-Hanif who worked as research assistants on this project, and Mez Khan for advising on the interpretation of the findings. We would also like to thank all of the research participants who shared their experiences with us. We would also like to thank Different Strokes for promoting this project.

## Statements and Declarations

### Author contributions

Xu Wang and Andrea Kusec designed the study. All authors contributed to the data collection, analysis, and writing of this manuscript. All authors have read and approved the final version.

### Ethics approval

Consent to participate and consent to publish: All participants provided informed consent to take part in this study and for their anonymised data to be published. Research ethics was approved by Leeds Beckett University (No. 119043)

