## Supplementary Materials for "Experiences of health care from stroke survivors and caregivers from minoritised ethnicities"

**Appendix 1:** Participant Interview Script

Hi, my name is [the interviewer’s name]. I would like to thank you for taking the time to discuss your experience of taking part in this research.

We are conducting this study because, based on research evidence, people from minority backgrounds, such as South Asian and Black individuals, are unfortunately more likely to have a stroke compared to White people and at a younger age. Because of this, it is important to understand experiences specifically from stroke survivors from minority backgrounds.

1. Can you tell us about your experience about the stroke care you received while you were in hospital?

- Prompt if needed: The services could include mental health or psychology care, speech language therapy, occupational therapy, physiotherapy, dietitian, healthcare workers, nurses, doctors etc.
- Prompt if needed: The treatment can relate to physical changes (for example, mobility, fatigue, using limbs, vision); thinking difficulties (e.g., language, understanding people, attention, memory); or other treatments/help (e.g., returning to work, emotional difficulties); and interpreters, assessment and therapy for other languages
- Prompt if needed: How well-supported did you feel by staff in general in the hospital (acute and/or rehabilitation ward)?

2. Looking back on your experience receiving care for your stroke in hospital:

- Was there time where you felt that your cultural, religious, or dietary needs were asked about and incorporated into your stroke care plan?
  - If yes: Can you tell me more about this?
  - If no: Can you tell me why not?

3. Were there moments that make you realise (think of) your own ethnicity/race when you were in hospital for your stroke?

- Prompt if needed: If yes, can you elaborate a bit more what happened (specifically about positive/negative realisations of ethnicity)
- Prompt if needed: If no, can you tell us a bit of why?

4. Can you tell us about your experience of accessing and receiving stroke care services from after you were discharged from the hospital until now?

- Prompt if needed: For example, stroke services you received at your home (visiting from OTs/Physio Therapists/SLTs, nurses, mental health/talking therapy, social services) and/or in the communities (e.g., early supported discharge, stroke care from your GP)?
- What about accessing and receiving stroke care and support from relevant charities (e.g., stroke association, different strokes, aphasia support) or private care?

5. Looking back on your experience receiving care for your stroke while you were at home:

- Was there a time where you felt that your cultural, religious, or dietary needs were asked about and incorporated into your stroke care plan?
  - If yes: Can you tell me more about this?
  - If no: can you tell me why not?

6. Were there moments that make you realise (think of) your own ethnicity/race when accessing/receiving stroke care after leaving hospital until now?

- If yes, can you elaborate a bit more what happened (specifically about positive/negative realisations of ethnicity)
- If no, can you tell us a bit of why?

7. To what extent did the professionals consider and ask about your culture and ethnicity and adapt your stroke care plan (if relevant)?

- Prompt if needed: For example, education about different risks of having a stroke for people from minority backgrounds?
- Prompt if needed: For example, food and drink modification, accessing a health care professional that spoke your native language, involving being part of your religious/spiritual community as part of your stroke care plan, celebration of cultural festivals
- Prompt: if they are showing you pictures, are they relevant to your culture?

9. Overall, have you ever felt that your experiences with professions after stroke were affected by your ethnicity/cultural background?

10. Do you have suggestions to make stroke treatment/service more accessible and/or suitable to survivors/carers from minoritized ethnic backgrounds such as Black and South Asian individuals?

11. Do you have anything to add or any questions?

**Supplementary Table 1**. Additional example quotations for themes and subthemes

| **Themes** | **Subthemes** | **Quotations** |
| --- | --- | --- |
| I feel different in stroke care | Not fitting the typical stroke picture | "*I remember I went to some of the, uh, organisations, uh, that, uh, basically support for, umm, stroke survivors. And when I go to there, I was, I felt a bit alone. Maybe it would have been helpful to just, you know, if you, do a support group with the people that have different backgrounds as well because, the one that I went to was specifically all of the people were, uh, British, and it was a bit tough […] but I felt different from those people. Umm, so I think it would have been helpful to create a specific support group for the people as well, just, you know, if they want to, just, uh, have different ethnical backgrounds as well*." (P7, Stroke survivor, Turkish)  "[*The peer support groups were] white people, themselves, and therapist was white people. Not, not anybody else giving therapy, but they speak only in English language*." (P10, Stroke survivor, Asian Indian) |
|  | Level of familiarity with British culture | "*I think, because of the fact that I can communicate, and I could communicate well, um, and I don’t have, I have a British accent, so I don’t think it [the care] was an issue for me*." (P23, Stroke survivor, Asian Indian) |
| Culturally inclusive stroke care is important but inconsistent | Not enough cultural humility in standard stroke care | “*They don't say, “Well, this girl is Afro-Caribbean, and she needs this. She needs that”. No, definitely no.t*” (P6, Stroke survivor, Mixed White & Black Caribbean) |
|  | Perceived cultural closeness builds trust | "*For example, some, some nurses and staff were, uh, actually from the ethnic minorities, were more, how do I, more considerate in some, some aspects and more empathetic…because obviously, they understood that, the culture and everything, so yeah*." (P15, Carer, Asian Pakistani)  "*In the hospital, I feel people were very respectful. And maybe that's also due to the fact that a lot of hospital staff are normally from minoritised backgrounds, like a lot of nurses are um, South Indian, or from the Philippines”* (P8, Stroke survivor, Asian Indian)  "*I think it felt a lot more, more you know, more comfortable, and you know you felt a lot more welcome. You know that feeling of being, being handled by somebody you trust and you feel comfortable. It’s this race thing is you know different with them being aged you know so, they have a lot of resentment, especially if they don’t find that connection. With people especially from other races*.” (P1, Carer, Black African/Black British) |
|  | Whose responsibility is it to start the conversation about cultural and religious needs? | “*If you're asking, did anyone ask me if I had any needs, nobody asked the question for then me to say “oh, thank you, I'm OK” or “thank you, I, I need ABC.” Um, no, that didn’t, didn't come up. I wouldn't say that was a problem. However, I think it would be a welcome introduction because obviously I'm sure that there are people who do have specific religious or other, you know, beliefs or needs*.” (P24, Stroke survivor, White and Black Caribbean)  “*I think professionals need to pay more attention to erm specificity in terms of people’s choices and the grounds and I think it plays a lot in people’s care. I think healthcare for people, for some people is not just, it's not just about the drugs, it’s not just about, it's not just about the healthcare in itself, I think er a whole lot of things contribute to people’s healthcare and I think cultural grounds are very important.*” (P2, Carer, Black African/Black British) |
| Personal interpretations of the role of ethnicity in care | Questioning whether ethnicity related to care | "*Overall, uh, there was, yeah, there was some empty parts. But I think the issues that I faced was not solely based on my ethnicity."* (P7, Stroke survivor, Turkish)  "*Sometimes the recipient isn't aware of what could have been available or what could have been asked, um, or what could have been offered, and didn't even realise, let's just say they didn't realise they, they were being discriminated against, or, they didn't realise that there was a, there was a prayer room, you know, whatever it may be, right, it just didn't come up. So they never thought of themselves as having received substandard care. Uhm, that may be the case. I can't say for certain. Um, equally, I can't say with any certainty whether anybody treated me any differently due to my race or perceived race”* (P24, Stroke survivor, White and Black Caribbean) |
|  | Attributing difficulties in care to stroke symptoms | "*It is difficult, cause, if you look at ah, stroke…it is difficult. Because, maybe, they couldn't talk to me because I, I couldn't talk*." (P17, Stroke survivor, Black African/Black British)  “The challenge would be, in a sense, my mother couldn’t speak for herself. Her speech wasn’t really there, and even now, because of the stroke, know what it is that she would want to say is, is difficult.” (P022, Caregiver, Black Caribbean) |
|  | Feelings of equity relate to having no cultural needs | *"I don't think my care, and, and then if I think of it from a non-race, and just um from my sort of background, I don't think I got, I think from an age or anything like that I think I got the care that I needed, based on what I needed*. " (P23, Stroke survivor, Asian Indian)  "I don't feel that we are isolated or being singled out as being the minority at all. And in fact, I think there, yeah, they treat us quite equally as other patients." (P013, Caregiver, Asian Chinese) |
| Families champion survivors' cultural needs, what about communities? | Families are advocates for stroke survivors’ cultural needs | "*We, I know that I had to, whenever it, it became a priority in my mind. So maybe day one, it wasn't a priority, but it was on day three. And then day four, my priority was on washing, bathing, cleaning and then maybe day seven, we were trying to recall, revisit chaplaincy services again, you know, so I was on top of that for all of this diary management of needs. But why were the healthcare assistances not? Why were nurses not?*" (P14, Carer, Asian Pakistani)  “*Erm well, I think for example, I think in the kind of condition that is stroke, I don’t think a Black person would trust any other person aside a family member to take care of his family needs*” (P1, Carer, Black African/Black British) |
|  | Opening up conversations about stroke in minoritised ethnicity communities | “*I have got some awareness of this issue, in the Black and South Asian community, there are a couple of things at play. Scepticism about medicine, lack of knowledge about health, healthcare and medical advancements in innovation...."* (P24, Stroke survivor, Mixed White & Black Caribbean)  "*Within South Asian communities, there's so much stigma towards mental health and disability*." (P8, Stroke survivor, Asian India)  “*I have no idea [why we do not discuss it]. If I knew, like you say, but you would have thought this, you know, it's the education. Like, I can, I can imagine the old people don't, don’t like to talk about [disability]. But the younger generations are the same*.” (P19, Stroke survivor, Asian Indian)  "*I don’t think a lot of these people [from ethnic communities] probably get seen to, until it’s too late. Um, so there might be a lot of people that have had stroke or a TIA and not known about it, especially at a younger age*." (P23, Stroke survivor, Asian Indian) |
|  | Needing to understand why there is a different risk of stroke | “*And secondly, also, I think erm we also want to be better inclusion in terms of family and you know looking at medical history and I think why is that? Because stroke is prevalent in certain families and then the families more likely to see the cause and you know we have to understand what is the culture of that family to that medical condition, that way you’re able to understand if there are need if there are need and phases that they need to start addressing and want to really go into the health plan*” (P1, Carer, Black African/Black British)  "*The other challenge to some extent is, trying to find out why it is that we are disproportionately represented. I mean, do we, do we typically have higher cholesterol levels than, than the median group*?" (P22, Carer, Black Caribbean)  *“I question why this has happened to me, because everyone’s like ‘well, it’s your race.’ And I’m like, right, so I know cholesterol should be higher, but, I don’t have the same diet as my parents. And as they, as they, the, the family, the family in [home country] for example, it’s completely different. My lifestyle is completely different. So, I’m still struggling to understand why, where does that factor in our make-up of our body? And when it’s so about, ‘well, you’re Black’ or it’s like, you know, I get sickle cell, like, I think I understand that a bit more, but, stroke, cholesterol, all of that, why is it such a big disparity between Asian, Black, ethnic, um, to the white person? Um, and especially if we’re all in the same country, and I think that’s what confuses me. I think I get it if you’re in different countries because that lifestyle, that food, that weather, you know, that, there, there’s so many different contributing factors to that, I get that. But when you’re in the same country and you’re born here. And if it wasn’t for the colour of my skin and my name, and you’d think, you know, I was a White person. So um, what is it in my make-up, my genes, that’s caused this?"* (P023, Stroke Survivor, Black Caribbean) |
